# Application of the Adaptive Validation Design to estimate the association between transmasculine/transfeminine status and self-inflicted injury among transgender and gender-nonconforming children and adolescents

**DOI:** 10.1101/2020.02.20.20024182

**Authors:** Lindsay J. Collin, Richard F. MacLehose, Thomas P. Ahern, Michael Goodman, Timothy L. Lash

## Abstract

**Background:** An internal validation substudy compares an imperfect measurement of a variable with a gold standard measurement in a subset of the study population. Validation data permit calculation of a bias-adjusted estimate, expected to equal the association that would have been observed had the gold standard measurement been available for the entire study population. Guidance on optimal sampling of participants to include in validation substudies has not considered monitoring validation data as they accrue. In this paper, we develop and apply the framework of Bayesian monitoring to determine when sufficient validation data have been collected to yield a bias-adjusted estimate of association with a prespecified level of precision.

**Methods:** We demonstrate the utility of this method using the Study of Transition, Outcomes and Gender—a cohort study of transgender and gender non-conforming children and adolescents. Transmasculine and transfeminine status were determined from the gender code in the electronic medical record at cohort enrollment. This status is known to be misclassified because it can indicate either gender identity or sex recorded at birth. Our interest is in the association between transmasculine and transfeminine status and self-inflicted injury. To address possible exposure misclassification, we demonstrate the method’s ability to determine when sufficient validation data have been collected to calculate a bias-adjusted estimate of association that is less than 80% greater than the precision of the conventional estimate.

**Results:** In the conventional age-adjusted analysis, we observed that transmasculine children and adolescents were 1.80-fold more likely to inflict self-harm than transfeminine youths (95%CI 1.27, 2.55). Using the adaptive validation approach, 200 cohort members were required for validation to yield a bias-adjusted estimate of OR=3.03 (95%CI 1.76, 5.56), which was similar to the bias-adjusted estimate using complete validation data (OR=2.63, 95%CI 1.67, 4.23).

**Conclusions:** Our method provides a novel approach to effective and efficient estimation of classification parameters as validation data accrue. This method can be applied within the context of any parent epidemiologic study design, and modified to meet alternative criteria given specific study or validation study objectives.

## Background

As gender identity develops in children, it may not match the sex recorded at birth.^1–3^ Although some transgender people reject binary gender definitions, a person whose gender identity differs from a male sex recorded at birth is often referred to as transfeminine, and a person whose gender identity differs from a female sex recorded at birth is often referred to as transmasculine.^4,5^ The development of gender identity and gender nonconformity in children and adolescents is an area of evolving research. Estimates suggest that 10% to 30% of gender nonconforming children may go on to identify with a gender that differs from their sex recorded at birth.^6^ Mental health conditions and their sequelae, such as self-inflicted injury, are an especially important concern for the health of transgender or gender nonconforming youth.^7–9^

The Study of Transition, Outcomes and Gender (STRONG) is an electronic medical record-based cohort study of transgender and gender nonconforming individuals which was established to understand long-term effects of hormone therapy and surgery on gender dysphoria, mental health, acute conditions such as injury, and chronic diseases such as cardiovascular disease and cancer.^10^ Cohort members were identified using International Classification of Disease-9 (ICD-9) codes that reported a diagnosis relating to gender dysphoria in the Kaiser Permanente healthcare database. Once cohort members were identified, transmasculine and transfeminine status were determined from the gender codes recorded in the electronic medical record. However, the gender codes recorded in the electronic medical record at the date of cohort entry sometimes reflected the person’s current gender identity and sometimes reflected the person’s sex recorded at birth. This uncertainty presented an important barrier to accurately classifying participants as transfeminine or transmasculine.

Given the importance of accurately classifying cohort members as transmasculine or transfeminine, implementation of an effective and efficient validation substudy design was essential to achieving the study’s aims. Guidance on optimal sampling of participants to include in a validation substudy pertain only to scenarios in which the complete parent study population has been enrolled and follow-up has been completed.^11,12^ Efficient designs for sampling participants as they are enrolled and followed have not been previously considered. Earlier work on designing validation substudies has also focused on setting the sample size required to reduce measurement error to a specified level or on optimizing the design given fixed resources available to support the validation study.^13–15^ They have not focused on optimizing the bias-adjusted estimate of effect.^16^ Given the cost of implementing a validation study, researchers may want to know at what point sufficient validation data have been collected to meet the objectives of validation.

Prospective monitoring of validation data as they accrue allows researchers to determine when sufficient validation data have been collected to obtain classification parameters that meet prespecified stopping criteria (for example, criteria that assure a precise bias-adjusted estimate of effect). Bayesian monitoring techniques have been used in clinical trials to estimate and monitor treatment response over time and to adapt the study design as data accrue, either by stopping the trial early or by modifying treatment allocation probabilities.^17^ Herein, we extend this conceptual framework to the development of an adaptive approach to validation study design. We use this new approach to calculate a bias-adjusted estimate of association for the association between transmasculine/transfeminine status and self-inflicted harm—accounting for possible misclassification of transmasculine/transfeminine status—with real-time validation data collection informed by a predefined stopping criterion for the precision of the bias-adjusted estimate of association between transgender status and the occurrence of self-inflicted injury. The STRONG cohort previously published elevated risks of self-inflicted injury and other mental health outcomes among transgender children and adolescents compared with population-based comparators.^18^ In this study, we developed and used the adaptive validation substudy design to calculate a bias-adjusted estimate of association for the association between transmasculine/transfeminine status and self-inflicted harm, accounting for the possible misclassification of transmasculine/transfeminine status. This strategy can be viewed as a sequential Bayesian analysis, in which the distributions of the sensitivity and specificity are estimated at specified intervals while the validation data accrue. At each time point, one uses the newly collected validation data to update the estimates of sensitivity and specificity, to compare the results against stopping criteria, and to decide whether to collect additional validation data. We demonstrate the utility of an adaptive validation design to sample validation data until the bias-adjusted estimate of association reaches a desired level of precision.

## Methods

### Study population

The STRONG youth cohort included individuals aged 3 through 17 at index date, identified using International Classification of Diseases 9^th^ Revision (ICD-9) codes and keywords related to transgender or gender non-conformity status in electronic medical records from Kaiser Permanente health plans in Georgia, Northern California, and Southern California. The index date corresponds to cohort entry and is defined as the first date with a recorded ICD-9 code or keyword reflecting transgender and gender non-conforming status between 2006 and 2014. Demographic data collected from the electronic medical records included the patient’s gender, but a person’s gender in the medical record could correspond to their gender identity or to their sex recorded at birth. This misclassification precluded accurate assignment of cohort members to transmasculine or transfeminine status based on their electronic medical record gender code. We will call this measurement the “misclassified sex recorded at birth.” To overcome this limitation of the available data, cohort members’ archived and complete medical records—the gold standard in this study—were reviewed to determine sex recorded at birth. Medical records were reviewed by keyword search in selected text strings to identify additional anatomy-related or therapy-related terms that would unambiguously indicate sex recorded at birth. We will call this the “gold standard sex recorded at birth.”

### Outcome

Self-inflicted injury was identified using ICD-9 codes, which included the date of occurrence of the first event. Self-inflicted injury was categorized as ever vs. never, which allowed for an event at any point over the course of cohort enrollment and follow-up due to the relatively small number of events. Median duration of follow-up was 10.5 years.

### Exposure status

The exposure of interest was transmasculine or transfeminine status, which can be determined from knowledge of the sex recorded at birth. The misclassified sex recorded at birth was based on the concurrent electronic medical record demographic data and known to be misclassified because it could either represent sex recorded at birth or concurrent gender identity. For example, if the cohort member had a gender code of ‘female,’ this code was assumed to refer to a ‘female’ sex recorded at birth, and therefore represented an individual who would identify as transmasculine. Similarly, a gender code of ‘male’ was assumed to refer to ‘male’ sex recorded at birth, and therefore represented an individual who would identify as transfeminine.

### Exposure validation

The misclassified sex recorded at birth variable was validated for members who were ≥18 years old as of January 1, 2015 (n=535; 40% of the youth cohort). Members under the age of 18 on this date were not validated, a design decision of the original investigators. For the quantitative bias analysis, we focus on sensitivity and specificity. Sensitivity was defined as the probability that sex recorded at birth in the concurrent medical record was female among those whose sex recorded at birth in the gold standard medical record was female. Specificity was defined as the probability that the sex recorded at birth in the concurrent medical record was male among those whose sex recorded at birth in the gold standard medical record was male. Our assignments of the labels “female” and “male” to sensitivity and specificity, respectively, were made at random.

### Adaptive validation sampling design

We used the STRONG cohort’s misclassified sex recorded at birth and the validation data on gold standard sex recorded at birth to calculate the classification parameters (sensitivity and specificity). Although the STRONG cohort had already completed enrollment, validation, and follow-up, we applied our Bayesian adaptive validation approach as if the study were accruing in real time. We determined the sample size necessary for estimates of sensitivity and specificity to meet our stopping criterion—a bias-adjusted estimate with precision no more than 80% wider than the conventional age-adjusted estimate, which did not account for exposure misclassification. Precision was measured by the ratio of the upper limit of the 95% interval to the lower limit of the 95% interval.

Among cohort members with validation data on their sex recorded at birth, we used an iterative beta-binomial Bayesian model to update the sensitivity and specificity.^19^ We assumed, before seeing any validation data, that all values of sensitivity and specificity were equally likely, due to the lack of prior information on recording gender identity in electronic medical records among children and adolescents. In settings where a literature exists, this information may be used to inform a prior distribution from previous validation studies. It would also be possible to correlate sensitivity and specificity through, for example, a receiver-operator characteristic curve. To simplify in this initial treatment, we kept the updating process for sensitivity and specificity independent, and to simplify notation, we refer to both parameters as θ:

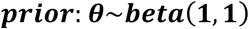

This beta distribution is identical to a uniform distribution, with all values of sensitivity and specificity having equal probability. The data in the validation study correspond to whether the i^th^ individual’s gold standard sex recorded at birth, y_i_, matched the gender in the concurrent medical record. That is, if a person’s complete medical record indicated that the person’s gold standard sex recorded at birth was male, then their validation data contributed to the estimate of specificity. If their observed misclassified sex recorded at birth was also male, then the validation result was concordant. If the person’s complete medical record indicated that the gold standard sex recorded at birth was female, then their validation data contributed to the estimate of sensitivity. If their observed misclassified sex recorded at birth was also female, then the validation result was concordant. The likelihood contributed by the i^th^ individual in the validation substudy is:

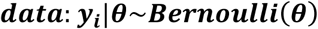

After the first individual’s validation data has been collected, the likelihood and prior can be combined via Bayes’ theorem, and the posterior distribution of the bias parameter (sensitivity or specificity) calculated as:

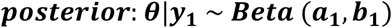

Where *a*_1_ = 1 + *y*_1_ and *b*_1_ = 2 − *y*_1_. The mean of this distribution can be used as an estimate of the bias parameter: 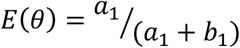. One could choose to use the median or mode of the distribution instead, and credible intervals can be defined using percentiles of the distribution. After the first observation has been accumulated, the posterior distribution becomes the prior distribution that is updated by the second observation, and this process repeats for each observation—or block of observations—collected:

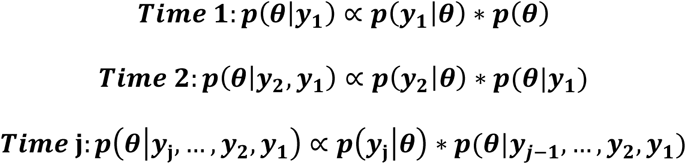

After the (J^th^) individual is validated, the posterior is:

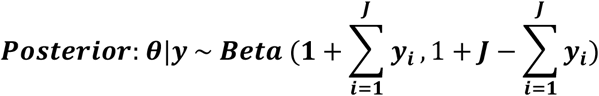

As above, the mean, median, mode, and credible intervals can be calculated from this distribution to give estimates of sensitivity or specificity at any point during the accumulation of validation data. We assume that any misclassification is nondifferential with respect to the outcome, which is defensible because the record of self-inflicted harm corresponded to an event chronologically after the record of sex recorded at birth (the gold standard) and the gender code (the misclassified variable). We updated sensitivity and specificity in blocks of ten, by taking a random sample of ten from each category of the gold standard sex recorded at birth. For each category, we randomly sampled ten individuals without replacement to inform the updated estimate of the bias parameters.

In a supplementary analysis, we additionally update the sensitivity and specificity classification parameters chronologically with cohort enrollment. We did this to illustrate how the sensitivity and specificity parameters change over time as each individual has their misclassified exposure information validated. In practice, this approach would most likely not be informative as it is usually more efficient to validate in blocks rather than single observations (*e*.*g*., some number of medical records reviewed in a day or some number of bioassays analyzed simultaneously).

### Quantitative bias analysis

We applied a stopping criterion for accrual of validation data based on the precision of the bias-adjusted estimate. We first calculated the conventional age-adjusted odds ratio (OR) and 95% confidence interval (CI) for the association between transmasculine/transfeminine and self-inflicted harm using logistic regression. Given that self-inflicted harm was rare, the logistic regression estimate provides a reasonable estimate of the risk ratio. We then employed probabilistic bias analysis for exposure misclassification using sensitivity and specificity estimates from multiple iterations of the adaptive validation approach.^20^ For each probabilistic bias analysis, we completed 1000 iterations and report estimates that account for information bias, with random error incorporated using bootstrap approximation. The validation and subsequent probabilistic bias analysis were continued until the precision of the bias-adjusted OR was no more than 80% greater than the precision of the conventional age-adjusted OR. All analyses were carried out in R (R Foundation, Vienna, Austria) and SAS v9.4 (Carey, NC).

## Results

The STRONG youth cohort included 1331 persons, of whom 710 (53%) were classified as transmasculine based on the misclassified sex recorded at birth code and 621 (47%) were classified as transfeminine. Enrolment into the study occurred between 2006 and 2014, with an overall study period of enrolment and follow-up of 10.5 years. In the study period, we identified 113 (16%) and 54 (8.7%) cases of self-inflicted harm among transmasculine and transfeminine individuals, respectively, using the potentially misclassified sex recorded at birth code (**Table 1**). In the age-adjusted model, the conventional odds ratio associating transmasculine status, versus transfeminine status, with self-inflicted harm was 1.80 (95% CI 1.27, 2.55), again using the potentially misclassified sex recorded at birth code to determine exposure status (**Table 2**). The precision of the conventional estimate of association was 2.01 (2.55/1.27).The youth cohort sensitivity and specificity of the misclassified sex recorded at birth code, calculated from complete validation, were 84% (95% CI 80%, 88%) and 90% (95% CI 85%, 93%), respectively. These were computed from a validated sample of 535 (40% of full cohort), including 283 (53%) transmasculine and 252 (47%) transfeminine cohort members who were >18 years of age as of January 1, 2015.

**Table 1:** Characteristics of the STRONG children and adolescent cohort population (n=1331). Transmasculine and transfeminine ascertained using the misclassified sex recorded at birth

| Characteristic | Transmasculine | Transfeminine |
| --- | --- | --- |
|  | n (%) | n (%) |
| **Total** | 710 (53) | 621 (47) |
| **Age, years** |  |  |
| 3–9 | 96 (14) | 155 (25) |
| 10–17 | 614 (86) | 466 (75) |
| **Self-inflicted Injury** |  |  |
| Yes | 113 (16) | 54 (8.7) |
| No | 597 (84) | 567 (91) |

**Table 2:** Conventional and bias-adjusted estimates of association between transmasculine/transfeminine children and adolescents and self-inflicted injury in the STRONG cohort.

| Method | Sensitivity | Specificity | Number Validated | OR (95%CI) | Precision |
| --- | --- | --- | --- | --- | --- |
| **Conventional** | 1.00 | 1.00 | 0 | 1.80 (1.27, 2.55) | 2.01 |
| **Adaptive Validation** | 0.81 (0.73, 0.88) | 0.90 (0.84, 0.95) | 200 | 3.03 (1.76, 5.56) | 3.16 |
| **Complete Validation Set** | 0.84 (0.80, 0.88) | 0.90 (0.85, 0.95) | 535 | 2.63 (1.67, 4.23) | 2.53 |

### Bias-adjusted estimates

Our pre-specified stopping criterion for the validation study was based on no more than an 80% increase in the ratio of the upper to lower confidence limits and precision of the bias-adjusted estimate, compared with the precision of the conventional estimate. We therefore continued with validation until the precision of the bias adjusted estimate was no greater than 1.8 × 2.01=3.62. This required a final validation sample of 200 cohort members, from which estimated values for sensitivity and specificity were 81% (95%CI 73%, 88%) and 90% (95%CI 84%, 95%), respectively (**Figure 1**). Application of the estimates of the bias parameters from the adaptive validation design yielded a final bias-adjusted estimate of OR=3.03 (95%CI 1.76, 5.56), incorporating both information bias and random error (**Table 2**). The bias-adjusted estimate obtained by using the complete validation data was OR=2.63 (95% CI 1.67, 4.23).

**Figure 1.**
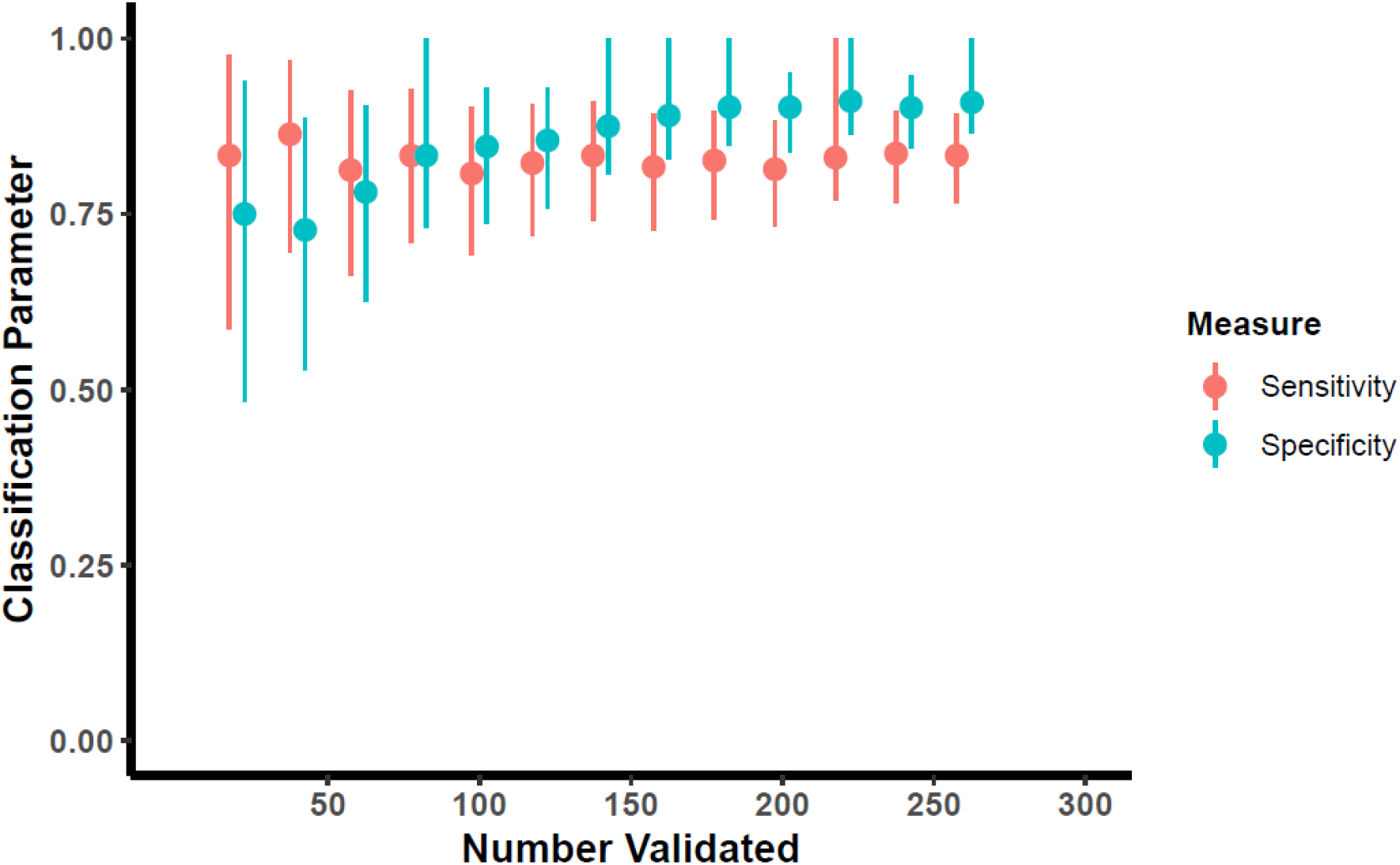
Adaptive validation using blocks of n=10 among STRONG children and adolescent cohort for estimation of sensitivity and specificity classification parameters.

## Discussion

In this study, we report that transmasculine children and adolescents are about three times more likely to inflict self-harm than transfeminine children and adolescents. The conventional estimate (OR=1.80; 95% CI 1.27, 2.55) obtained using the potentially misclassified gender code was substantially closer to the null than the bias-adjusted estimate of association obtained by accounting for misclassification of transmasculine/transfeminine status (OR=3.03; 95% CI 1.76, 5.56) using results from 200 participants included in the adaptive validation design. This estimate is similar to the estimate obtained using the complete validation data (OR=2.63; 95% CI 4.23, 1.67), which used all 535 validated records.

The STRONG youth cohort investigators previously reported that transgender and gender nonconforming children and adolescents were more likely to present with mental health conditions, especially those related to anxiety and depression, than population-based comparators.^18^ Furthermore, prevalence ratio estimates of self-inflicted harm were especially pronounced among both transmasculine and transfeminine individuals compared to population-based referents of the same age. The estimates were imprecise, however, due to sparse number of events among cisgender children and adolescents. Compared to reference males, transfeminine children and adolescents had 3.9 times the prevalence of self-inflicted injury (95% CI 1.8, 8.2). Similarly, compared to reference females, transmasculine children and adolescents had 8.7 times the prevalence of self-inflicted injury (95% CI 5.9, 12.8). In this study, we additionally report that transgender and gender nonconforming youths that are classified as transmasculine are more likely to inflict self-harm than those classified as transfeminine, consistent with the pattern of associations when transgender youth were compared to cisgender comparators.

The sensitivity and specificity estimates calculated from the adaptive validation approach were similar to those obtained from the complete validation substudy, though estimated with less precision. We observed that among transmasculine children and adolescents, the concurrent gender code was less likely to reflect the sex recorded at birth compared with those classified as transfeminine in the same age category. This finding is in agreement with the STRONG investigator’s previous observation that the proportion of transmasculine people who felt they were perceived by others as males was higher than the corresponding proportion of transfeminine people who felt they were perceived by others as females.^21^

An important advance is the development of an approach to validation study design suitable for scenarios in which validation data are collected in real time, and applicable to any parent epidemiologic study design. This method provides a valuable tool for prospective validation, allowing researchers to optimize fixed study resources when implementing validation studies. We demonstrated the ability of the method to determine when a validation study has generated sufficient data to support a prespecified precision of a bias-adjusted estimate of association. Using this design, the sensitivity and specificity calculated from the limited sample in the adaptive validation set were comparable to those obtained from full validation efforts. However, fewer persons required validation using the adaptive design.

Validation data are often expensive to collect. When the gold standard is measured by medical record review, substantial labor and data access costs accrue. When the gold standard is measured by bioassay, specimens must be collected, stored, and analyzed. It is almost inherently true that the gold standard measure is more expensive or difficult to collect than the misclassified measure; otherwise the gold standard would have been collected in the first place. Conventional designs for validation studies use fixed sample sizes or fixed resource allocation, without incorporating prior knowledge or considering validation data as they accrue. Using this adaptive validation design can therefore save substantial costs and time, which can then be allocated to other research objectives. Iterative updating of classification parameters informs values of the parameters of interest and can be used as a marker for when sufficient information has been collected, indicating when validation efforts can stop. This process allows researchers to decide, in real time, how to efficiently allocate resources to validation studies, with the ability to adapt based on the specific needs of their study. There is often a trade-off with internal validation studies—the possible sacrifice of sample size or other data collection of the parent study to obtain assurance that the variables of interest have adequate validity. Our method may enhance the ability of researchers to save on resources allocated to validation, by thoughtfully validating over the study time period while periodically assessing the performance of the variables mismeasured in the study. In this approach, the sample size necessary to achieve the specified stopping criterion is potentially smaller than would be required by other approaches, and validation can be completed in parallel with cohort enrollment.

This proposed validation study design is amenable to alternative stopping rules based on prior knowledge, other criteria, or other validation study goals. For example, researchers may prefer a precision-based stopping rule, such as a pre-specified width of the interval around the classification parameter estimates that must be achieved before stopping, which can be easily incorporated into this method. For all stopping rules, special consideration should be given to random error, which could inadvertently cause the validation study to stop too soon. Investigators may be interested in having stopping rules evaluated only after some minimum number of records have been validated, for instance.

In this study, we used the adaptive validation design to estimate sensitivity and specificity, as we had complete validation data for 40% of the study population and could condition on the true sex recorded at birth. However, a more conventional approach would be to condition on the observed exposure status, which would allow estimation of the positive and negative predictive values. A limitation of predictive values is that they are specific to the study from which they arise, and are not readily applied outside of the given study population. Sensitivity and specificity are more easily transportable to other populations, and therefore more amenable to starting the adaptive validation analyses with an informative prior. Positive and negative predictive values are dependent on the prevalence of the measure of interest, so researchers should calculate the positive and negative predictive values of exposure within strata of the outcome. However, this is difficult to accomplish in a study that prospectively validates data, as the outcome status may not be known at the time of validation. In validation studies that are initiated after the parent study is completed, this could be more easily accomplished. For studies that evaluate the association between an exposure and multiple outcomes, additional considerations will apply, such as validation of exposure within strata of each outcome. supplementary analysis, in which we updated the sensitivity and specificity contemporaneously with validation study enrollment, we observed a time-trend in the validity of exposure classification (**Figure 2**). The change in classification parameters over time may be due, in part, to the socio-political context surrounding transgender health. It is conceivable that such time trends could exist in other scenarios and measures that require validation. Note, however, that the time-trend provides potentially useful information that might easily be missed if all validation data were collected and analyzed after the primary data collection was complete. Detection of a time-trend may serve as its own stopping criterion, as the researchers would want to make sampling adjustments to conduct the adaptive validation approach within time periods that would more accurately capture the time-trend in classification parameters. Furthermore, the corresponding bias-adjustment would need to take the time-trend into account, improving the accuracy of bias-adjusted results. Other approaches do not allow for detection of a time-trend, as illustrated by the fact that the trend was not, in fact, previously noted in the STRONG cohort analyses.

**Figure 2:**
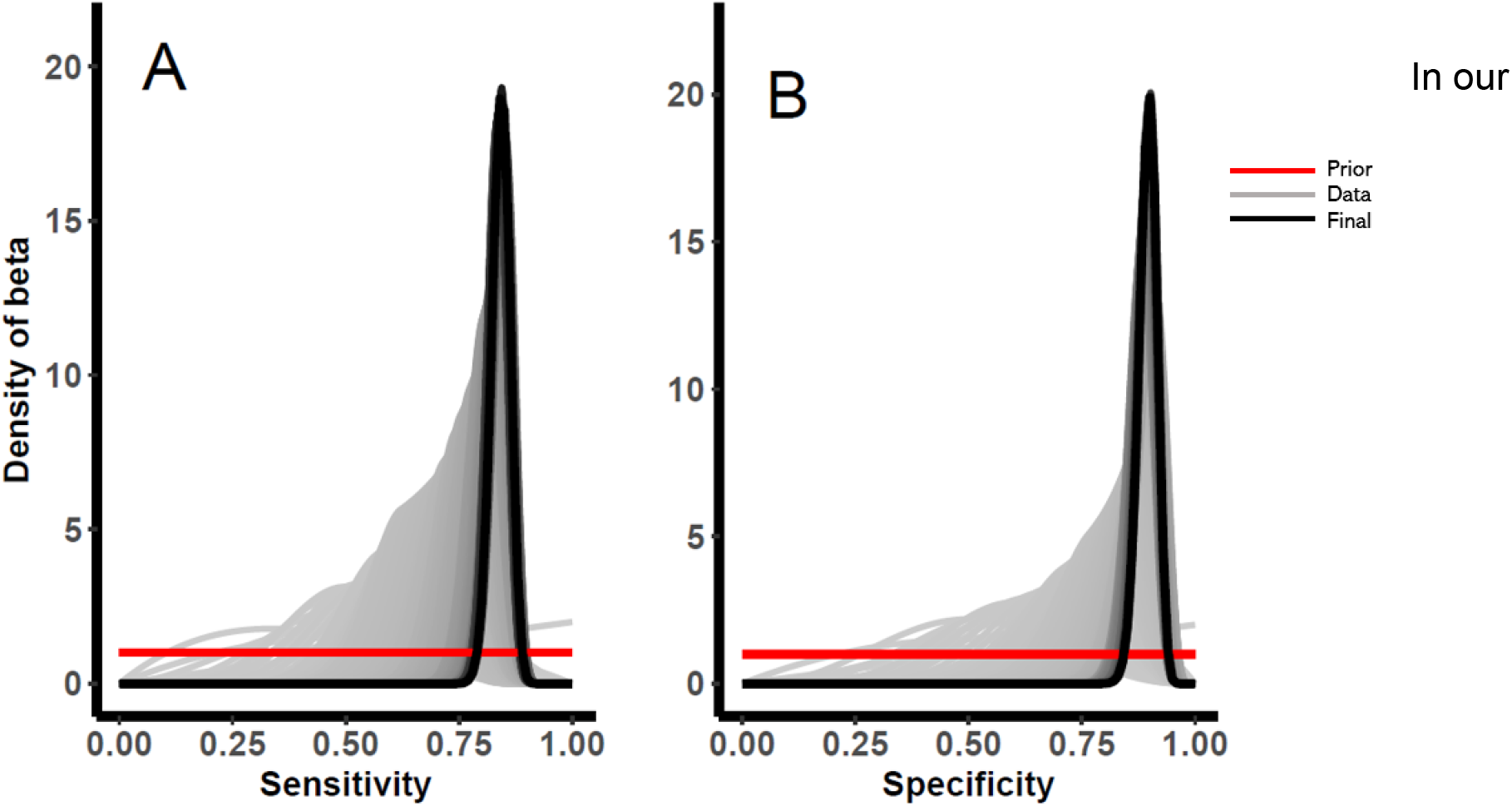
Adaptive validation design, updating sensitivity and specificity classification parameters as each individual is enrolled in the validation study among STRONG children and adolescents.

## Conclusions

Our proposed adaptive validation design is useful for calculating classification parameters as validation data accrue in epidemiologic studies, which can lead to effective and efficient conduct of validation substudies. Extending this proposed method to studies with multiple outcomes and for use with positive and negative predictive values are important goals for future development. In this application, the association between transmasculine status (versus transfeminine status) and self-harm in children and adolescents was substantially underestimated by the potentially misclassified gender code. The bias-adjusted estimate, made by applying internal validation data, yielded a stronger association, even when only part of the validation data were used.

## Data Availability

Due to patient confidentiality, data are only available upon IRB approval from the research institution in collaboration with Dr. Michael Goodman and the STRONG research team. Example code used to perform the adaptive validation study is available from GitHub (https://github.com/lcolli5/Adaptive-Validation).

## Notes

**Conflicts of Interest:** The authors declare no conflict of interest.

**Sources of Funding:** This work was supported in part by the US National Cancer Institute (F31CA239566) awarded to Lindsay J Collin, (R01CA234538) awarded to Timothy L Lash, and the US National Library of Medicine (R01LM013049) awarded to Timothy L Lash. Thomas P Ahern was supported by an award from the US National Institute of General Medical Sciences (P20 GM103644). STRONG cohort data were collected with support from Contract AD-12-11-4532 from the Patient Centered Outcome Research Institute and by the NICHD R21HD076387 awarded to Michael Goodman.

### Competing Interest Statement

The authors have declared no competing interest.

### Funding Statement

This work was supported in part by the US National Cancer Institute (F31CA239566) awarded to Lindsay J Collin, (R01CA234538) awarded to Timothy L Lash, and the US National Library of Medicine (R01LM013049) awarded to Timothy L Lash. Thomas P Ahern was supported by an award from the US National Institute of General Medical Sciences (P20 GM103644). STRONG cohort data were collected with support from Contract AD-12-11-4532 from the Patient Centered Outcome Research Institute and by the NICHD R21HD076387
awarded to Michael Goodman.

